# Predictors of SARS-CoV-2 anti-Spike IgG antibody levels following two COVID-19 vaccine doses among children and adults in the Canadian CHILD Cohort

**DOI:** 10.1101/2023.09.06.23294696

**Authors:** Rilwan Azeez, Larisa Lotoski, Geoffrey L. Winsor, Corey R. Arnold, Yannick Galipeau, Martin Pelchat, Stephanie Goguen, Elinor Simons, Theo J. Moraes, Piush J. Mandhane, Stuart E. Turvey, Shelly Bolotin, David M. Patrick, Jared Bullard, Lisa M. Lix, Natasha Doucas, Natalie Rodriguez, Fiona S.L. Brinkman, Padmaja Subbarao, Marc-André Langlois, Meghan B. Azad

## Abstract

**Background:** Vaccination helps prevent SARS-CoV-2 infection and severe COVID-19. However, vaccine-induced humoral immune responses vary among individuals and wane over time. We aimed to describe the SARS-CoV-2 anti-spike IgG antibody response to vaccination and identify health and demographic factors associated with this response among children and adults.

**Methods:** We studied a subset of double-vaccinated children (n= 151; mean age: 12 ±1.5 years, 46% female) and adults (n= 995; 44 ±6.0 years, 60% female) from the Canadian CHILD Cohort. Dried blood spots were collected over two time periods (March 2021 to September 2021; October 2021 to January 2022). Antibody levels were quantified using automated chemiluminescent ELISAs. Demographic, vaccination, and health data were collected via online questionnaires. Associations were determined using multivariable regression.

**Results:** Our cohort had SARS-CoV-2 anti-spike seropositivity rate of 97% following two COVID-19 vaccine doses. In both children and adults, the highest antibody levels were observed around three months post-vaccination and did not differ by biological sex. Higher antibody levels were associated with: prior SARS-CoV-2 infection (β=0.15 scaled luminescence units, 95%CI, 0.06-0.24), age <18 years (β=0.15, 95%CI 0.05-0.26) and receiving the Moderna mRNA (β=0.23, 95%CI 0.11-0.34) or Pfizer-BioNTech mRNA vaccines (β= 0.10, 95%CI, 0.02-0.18) vs. a combination of mRNA and Oxford-AstraZeneca viral vector vaccines. There were no differences in antibody levels when comparing a 3-8 vs. 9-16-week interval between vaccine doses.

**Interpretation:** We identified key factors associated with post-vaccination antibody responses in children and adults, which could help improve future vaccine development and deployment among different population subgroups.

## Introduction

Vaccination is the most effective way to prevent SARS-CoV-2 infection and severe COVID-19^1,2^. However, vaccine-induced humoral immune antibody responses vary among individuals and wane over time^3^. Pre-existing health and sociodemographic factors have been associated with the varied immune response to vaccination^3^. Age, biological sex, prior SARS-CoV-2 infection, vaccine type and dosing intervals have all been reported to influence the magnitude of vaccine-induced immune responses among different population subgroups^4–7^. However, the variation and timing of this immune response is not fully understood, particularly in children. Further, few COVID-19 vaccine studies have involved participants with heterogeneous (“mix-and-match”) vaccine combinations. Understanding the determinants of COVID-19 vaccine responsiveness is an important research priority that could provide more insights on ways to improve COVID-19 vaccine immunogenicity and efficacy among different population subgroups.

In this study, we described the SARS-CoV-2 anti-spike IgG antibody response to vaccination and identified health and demographic factors associated with vaccine-induced antibody response among double-vaccinated children and adults in the Canadian CHILD Cohort.

## Methods

### Study design and population

We conducted a SARS-CoV-2 anti-spike and anti-nucleocapsid seroprevalence study using dried blood spots collected in two phases (Phase A: March 2021-September 2021; Phase B: October 2021-January 2022) among participants from the CHILD COVID-19 Add-on Study, embedded within the Canadian CHILD Cohort Study. Eligibility, consent, recruitment, and detailed methodology are as described elsewhere^8^. Notably, Phase B collection started one month before the approval of vaccines for children aged 5-11 years in Canada. In total, 3059 participants (1501 children and 1558 adults) contributed serology data for the current analyses (**Figure 1**). Participants provided one (n=1242) or two (n=1817) samples (n=4876 samples total); all but 58 Phase B participants contributed samples in Phase A. For analyses focused on double vaccinated participants, we limited our subsample to those who received two COVID-19 vaccine doses prior to their first or second DBS sampling (151 children and 995 adults).

**Figure 1.**
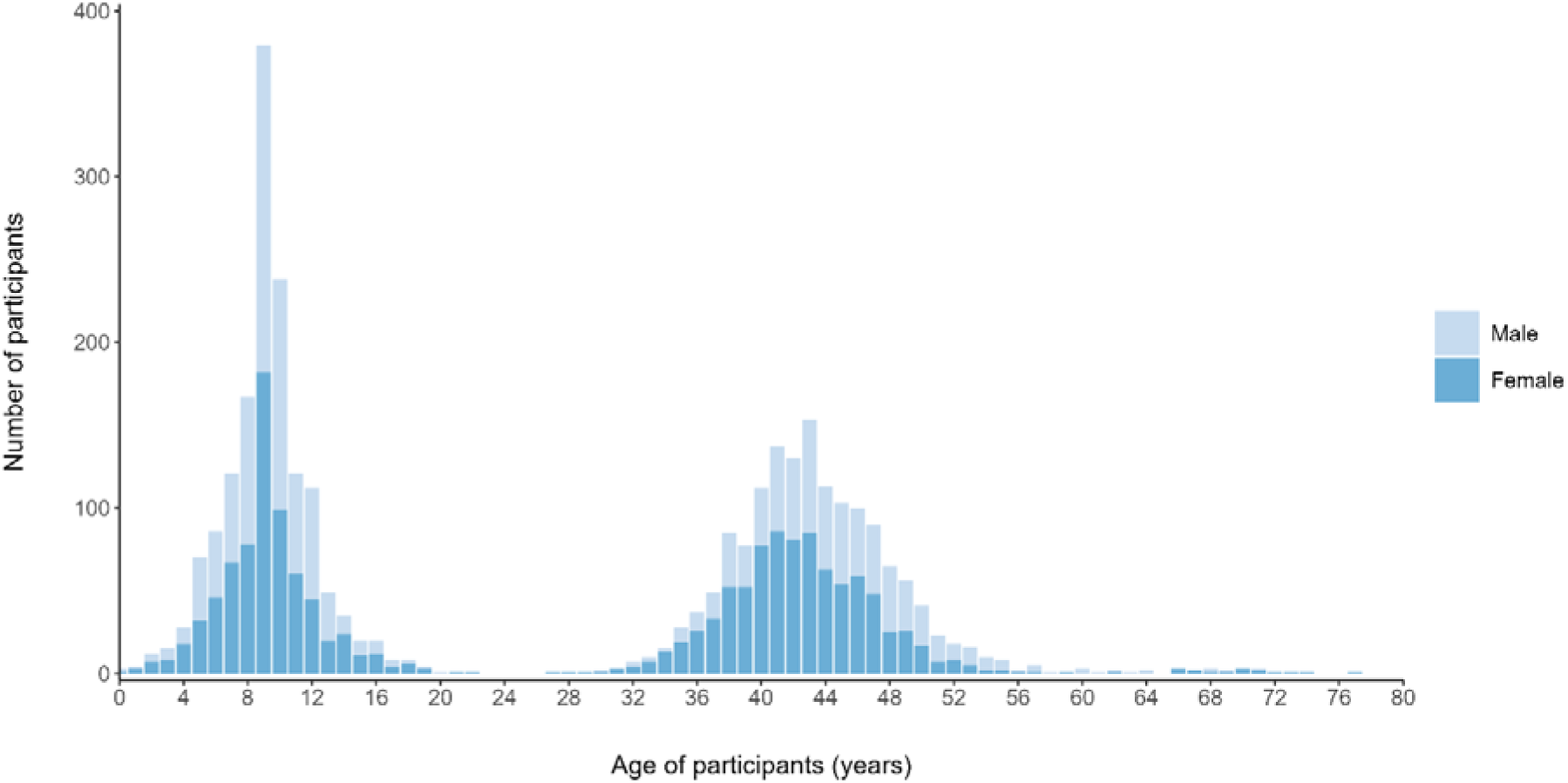
Age and biological sex of participants contributing serology data in the CHILD COVID-19 Add-On Study. N=3059 individuals (1501 children and 1558 adults).

### Data collection

Quarterly questionnaires collecting information were delivered using REDCap^9,10^ at baseline (January to June 2021) and three follow-up periods (August to September 2021, October to December 2021, January to March 2022) to capture: demographics, employment, COVID-19 testing, diagnosis, and vaccination uptake, and comorbidities. Brief biweekly questionnaires were deployed to capture COVID-19 testing and diagnosis. Blood for serology was collected from March 2021 to September 2021 (Phase A) and October 2021 to January 2022 (Phase B) using Dried Blood Spot (DBS) kits delivered and returned by mail.^11–13^

### COVID-19 case definition

#### Self-reported SARS-CoV-2 infection measures

Self-reported infection was captured through the biweekly and quarterly questionnaires. Participants were considered to have SARS-CoV-2 infection after reporting a positive COVID-19 test ascertained through any of the following: polymerase chain reaction test, mouth rinse test, or antibody/serology test.

#### SARS-CoV-2 serology

We measured SARS-CoV-2 anti-spike and anti-nucleocapsid immunoglobulin G (IgG) antibodies on automated chemiluminescent enzyme-linked immunosorbent assays (ELISAs) at the University of Ottawa Serology and Diagnostics High Throughput Facility as previously described.^14^ Participants with both anti-spike and anti-nucleocapsid levels above the pre-determined cut-off of 0.16 units^14^ were considered seropositive due to previous infection.

### Statistical analysis

Anti-spike and anti-nucleocapsid dried blood spot serology levels had skewed distributions and were compared using the Mann–Whitney test for two groups or Kruskal Wallis test for more than two groups. Among double-vaccinated participants, we used a) polynomial linear regression models with locally estimated scatterplot smoothing^15^ (LOESS) to visualize trends in antibody levels by time since second vaccination, and b) univariable and multivariable linear regression analyses to identify predictors of antibody levels. Candidate predictors were selected based on domain expertise and factors known to be associated with vaccine humoral responses. All covariates with *p*-values <0.2 in univariable models were included in a multivariable model to assess the independent relationships with anti-spike IgG antibodies. Results are reported as beta-estimates and 95% confidence intervals (95% CIs).

## Results

The mean age of participants was 44 (SD ±6) years among adults and 12 (SD ±1.5) years among children **(Figure 1)**. The majority identified as having European ancestral origins (60%) and most adults (75%) had a university degree **(Table 1)**. About 45% of families had an essential worker (defined as working in healthcare, delivery, retail, security or building maintenance) in their household. Notably, children (<18 years) are under-represented in the double-vaccinated serology sub-cohort (13% compared to 52% of the overall cohort) because vaccines were approved later for this age group.

**Table 1.**
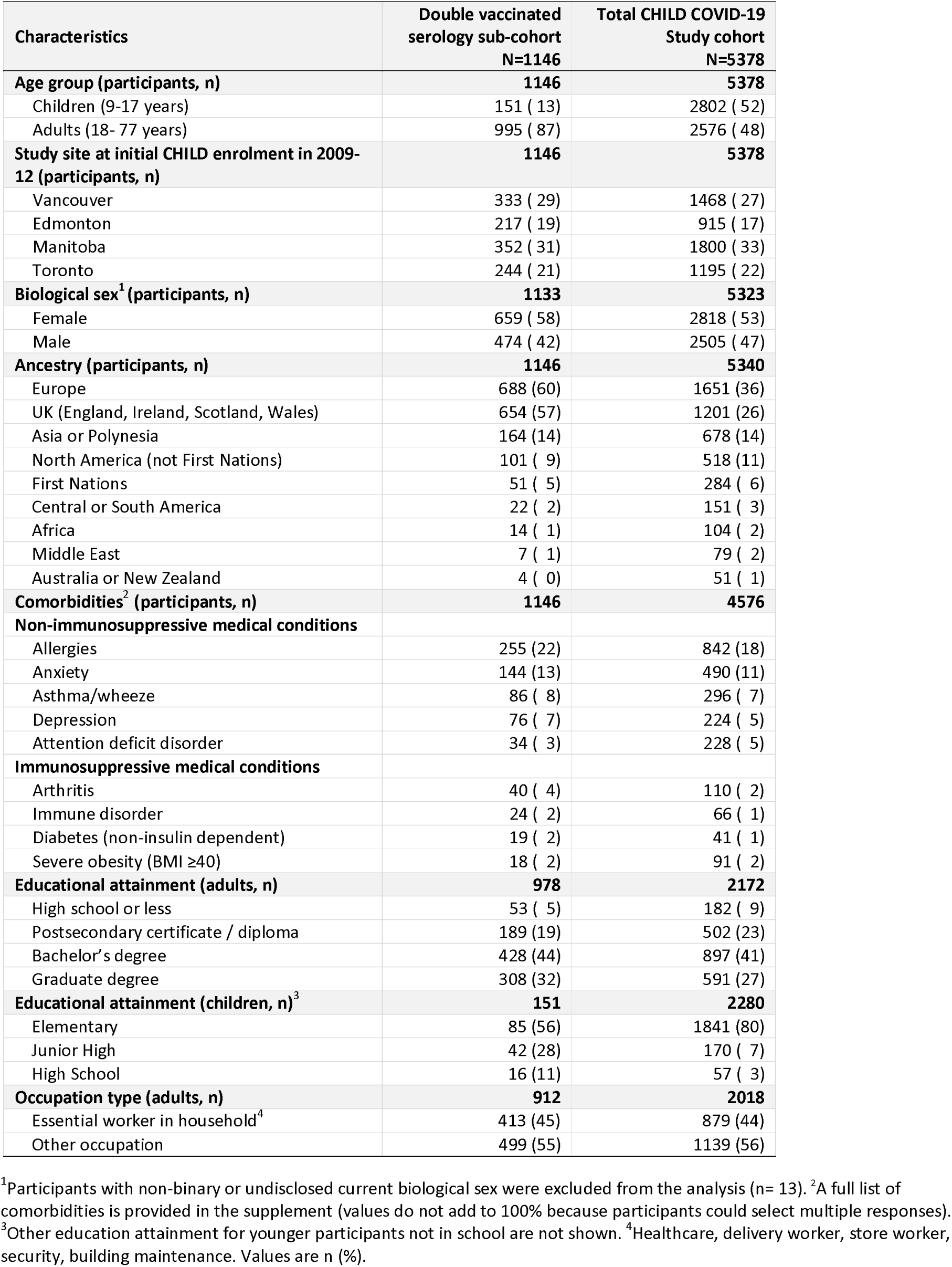
Baseline demographic characteristics of CHILD COVID-19 Study participants: overall and restricted to those with two vaccine doses prior to serologic testing.

Vaccine types and timing for double-vaccinated participants are summarized in **Table 2**. Among adults, 66% received mRNA vaccines for both doses (47% BNT162b2, 11% mRNA-1273, 8% combination of the two), while 30% received a combination of viral vector (ChAdOx1-S) and mRNA vaccine, and 4% received viral vector vaccines only. Children almost exclusively (99%) received the BNT162b2 mRNA vaccine for both doses. Participants received their second dose a median of 57 (IQR 41-65) days after the first dose. The mean interval between receipt of second vaccine dose and DBS sampling was 103 days (SD± 37) for children and 133 days (SD± 55) for adults.

**Table 2.** Vaccine types and time intervals among double-vaccinated participants.

| **Combination for both vaccine doses** | **Adults**<br>**N=995** | **Children**<br>**N=151** |
| --- | --- | --- |
| **Vaccine combination, n (%)** |  |  |
| mRNA BNT162b2 (Pfizer BioNTech) | 470 (47) | 149 (99) |
| mRNA-1273 / BNT162b2 + ChAdOx1-S | 296 (30) | 0 ( 0) |
| mRNA-1273 (Moderna) | 111 (11) | 0 ( 0) |
| mRNA-1273 + BNT162b2 | 80 ( 8) | 1 ( 1) |
| ChAdOx1-S (Oxford-AstraZeneca) | 34 ( 4) | 0 ( 0) |
| **Days between first and second dose, median (IQR)** | 57 (41-65) | 45 (33-54) |
| **Days since second dose, mean ± standard deviation** | 133 ± 55 | 103 ± 37 |

IQR, interquartile range.

### Anti-spike SARS-CoV-2 IgG antibody response following vaccination

Median anti-spike IgG antibody levels were significantly higher in individuals with two COVID-19 vaccine doses (1.81 scaled luminescence units, IQR 1.55-2.14) compared to unvaccinated (0.08, IQR 0.05-0.10; *p* <0.001), or single vaccinated participants (1.76 units, IQR 1.37-2.10 *p* <0.001), but lower than those who had received a third vaccine dose (1.90 units, IQR 1.67-2.24, *p* <0.001). Among double-vaccinated participants, antibody responses were highly variable, ranging nearly 3 orders of magnitude from 0.02 to 3.02 scaled luminescence units **(Figure 2A)**. About 3% had antibody levels below the seropositivity threshold and were classified as “non-responders” (children: 3%, n=5/151; adults: 3%, n= 26/995).

**Figure 2.**
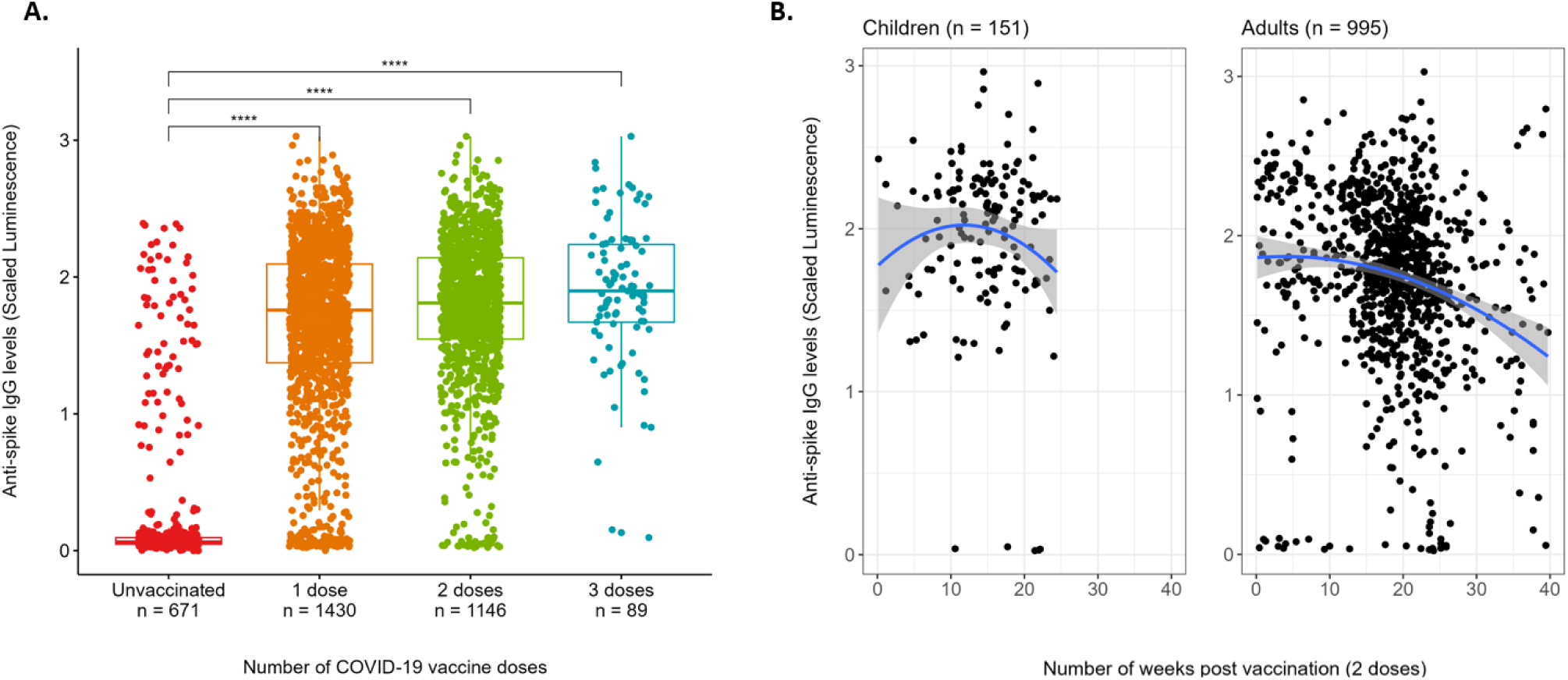
Anti-spike SARS-CoV-2 IgG antibody levels according to number of COVID-19 vaccine doses (A) and time since vaccination (B) in the CHILD Cohort. Serology from dried blood spots collected between March 2021 and January 2022. A) Horizontal lines indicate medians. B) Curved blue lines and gray shaded bands represent fit and 95% confidence interval from a polynomial linear regression model. Time range is larger for adults because vaccines were approved later for children. ****Kruskal-Wallis *p*-value <0.001.

### Vaccine responses by age, sex, vaccine type and comorbidities

Among double-vaccinated participants, children had significantly higher median antibody levels (2.08 units, IQR 1.71-2.24) than adults (1.79 units, IQR 1.49 -2.09, *p* <0.001) (**Figure 3A**). For both age groups, the highest antibody levels were observed approximately three months (thirteen weeks) after the second dose (**Figure 2B**), and no sex differences were observed (*p* =0.42) **(Figure 3B)**.

**Figure 3.**
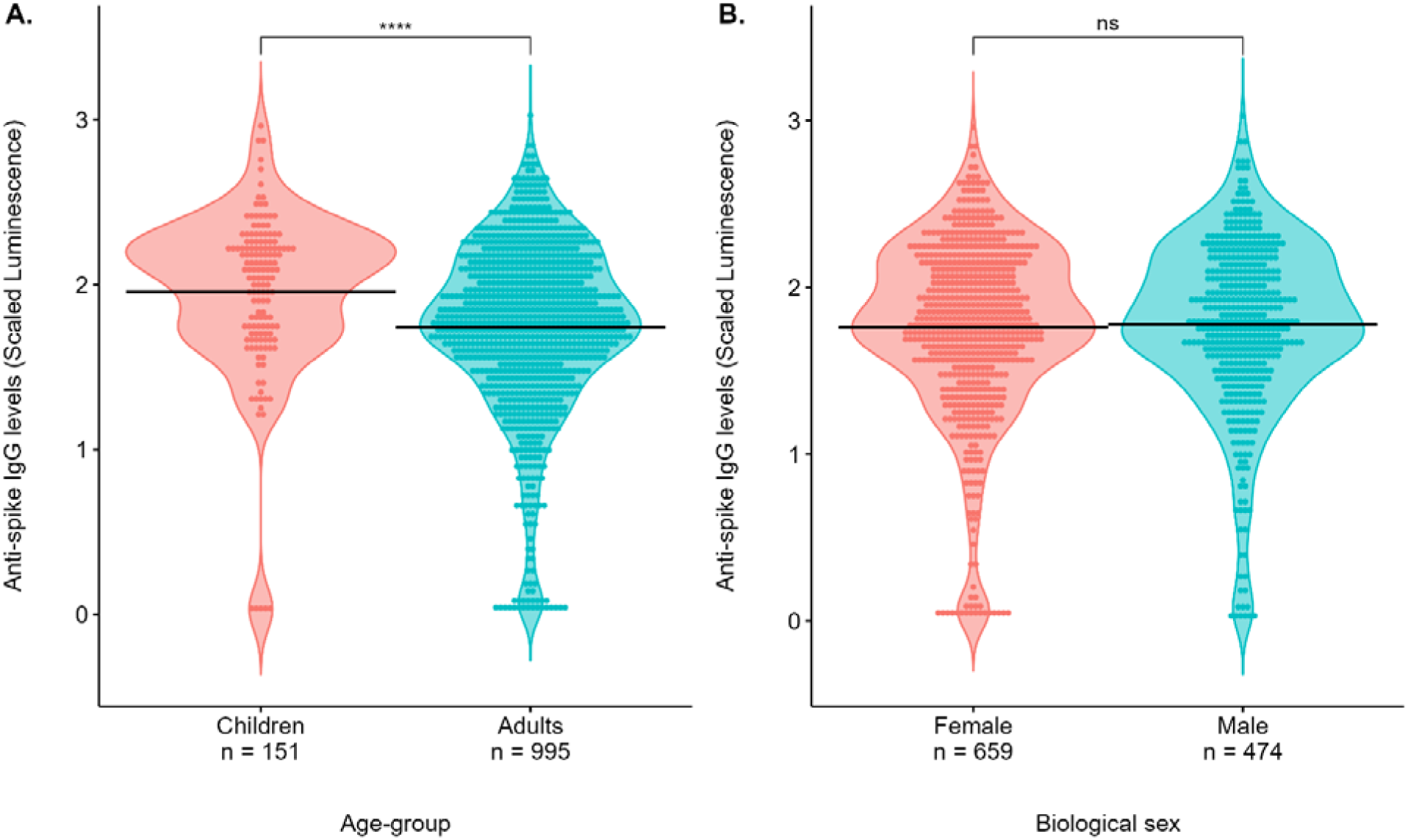
Anti-spike SARS-CoV-2 IgG antibody levels following two COVID-19 vaccine doses by age and sex in the CHILD Cohort. Serology from dried blood spots collected between March 2021 and January 2022. Horizontal lines indicate medians. ****Mann Whitney U p-value <0.001; ns - not significant.

Over half of double-vaccinated participants received mRNA vaccines for both vaccine doses (n= 811/1146, 99% of children and 66% of adults). Participants receiving mRNA-1273 or mRNA-BNT162b2 for both vaccine doses had significantly higher median antibody levels (1.96 units, IQR 1.75-2.18) and (1.85 units, IQR 1.57-2.18), respectively, in comparison to viral vector ChAdOx1-S vaccinated participants (1.16 units, IQR 0.65-1.81) (*p* <0.001) (**Figure 4A**). These findings were also observed across measured time-points in a polynomial linear regression model (**Figure 4B**).

**Figure 4.**
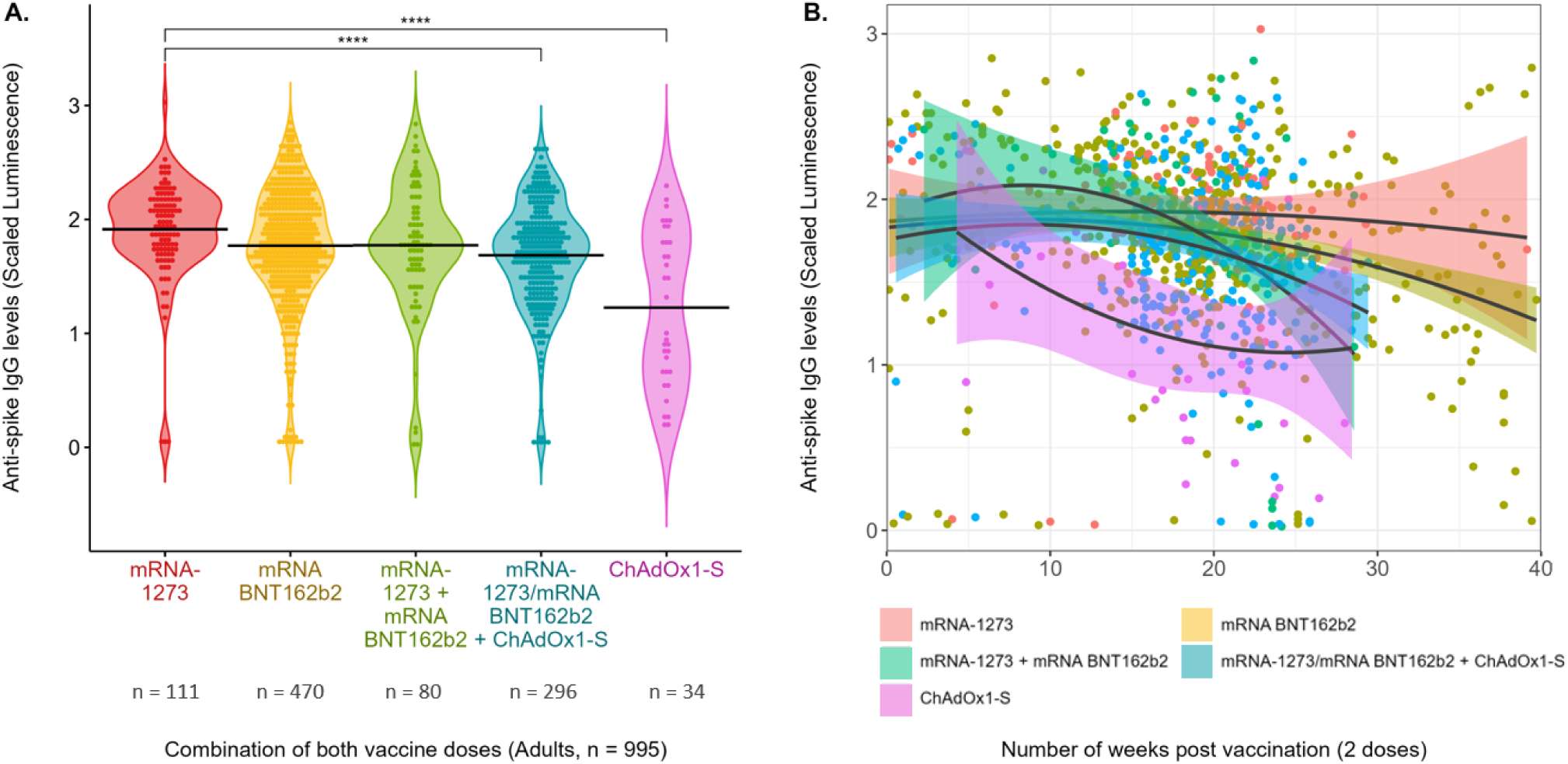
Anti-spike SARS-CoV-2 IgG antibody levels by type of COVID-19 vaccine received among double-vaccinated adults in the CHILD Cohort. Cross-sectional serology data from dried blood spots collected between March 2021 and January 2022. **A)** Horizontal lines indicate medians. **B)** Curved lines and shaded colored bands represent fit and 95% confidence interval from a polynomial linear regression model. ****Kruskal-Wallis *p*-value <0.001.

The most common medical conditions among participants were allergies, anxiety, asthma, and depression (range: 4-22%) **(Tables 1 and S1**); none of which were associated with vaccine responses. We had limited power to evaluate immunosuppressive conditions (defined as cancer, insulin and non-insulin dependent diabetes, inflammatory bowel disease, kidney disease, pneumonia, and severe obesity) as these affected less than 5% of participants.

### Antibody levels and vaccine responses in previously infected and infection-naïve participants

About 13% (n=147/1146) of double-vaccinated participants had evidence of prior COVID-19 when considering both serology and self-report. Prevalence was higher among adults (14%, n= 138/995) than children (6%, n= 9/151) (*p*= 0.007) but did not differ by sex in either age group (*p*= 0.42 for children; *p*= 0.43 for adults) (**Table 3**). Hybrid immunity (elicited through a combination of infection and vaccination^16^) was associated with higher and more rapid production of antibodies **(Figure 5A)**. Following receipt of two COVID-19 vaccine doses, antibody levels peaked within the first month among previously infected participants, compared to four months post-vaccination among infection-naïve individuals **(Figure 5B)**.

**Figure 5.**
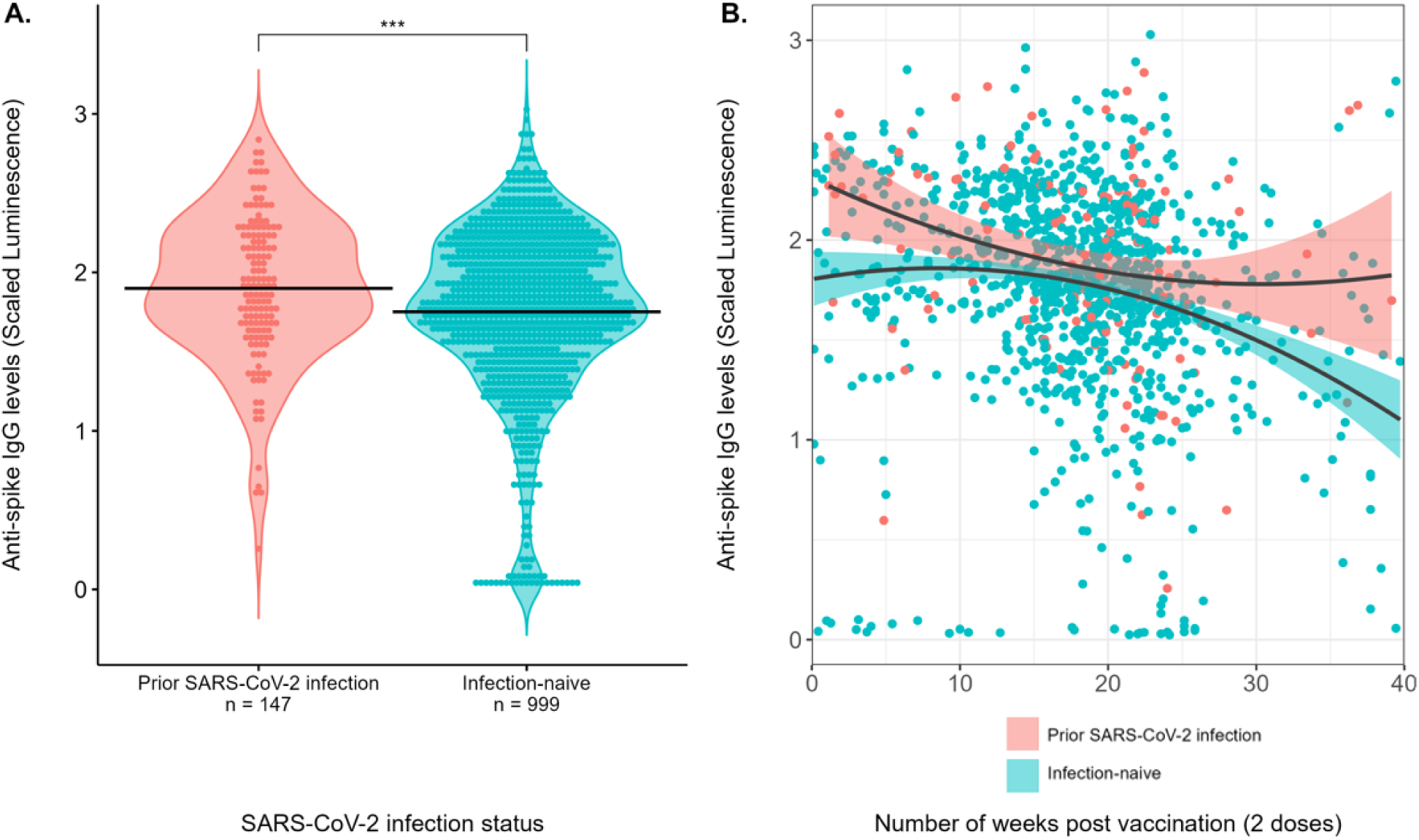
Anti-spike SARS-CoV-2 IgG antibody by SARS-CoV-2 infection status (based on self-report and serology) among double-vaccinated children and adults in the CHILD Cohort. Serology from dried blood spots collected between March 2021 and January 2022. **A)** Horizontal lines indicate medians. **B)** Hybrid immunity (combined infection and vaccination) appears to increase and slow the decay of antibody levels. ***Mann Whitney U *p*-value =0.001.

**Table 3.** SARS-CoV-2 infection prevalence among double vaccinated participants, by age group and sex.

| **Double-vaccinated participants** | **Prior SARS-CoV-2 infection, n (%)** | **Chi-square *p*-value** |
| --- | --- | --- |
| **Age-group, n (%)** |  |  |
| Children | 9/151 (6.0) | 0.01 |
| Adults | 138/995 (13.9) |  |
| **Biological sex<sup>1</sup>, n (%)** |  |  |
| Female | 90/659 (15.0) | 0.47 |
| Male | 57/474 (12.6) |  |

Values are n (%) for non-missing data for each variable. <sup>1</sup>Participants with non-binary or undisclosed current biological sexes were excluded from the analysis due to low statistical power (n= 13).

### Independent predictors of antibody levels in double-vaccinated individuals

In multivariable analysis **(Figure 6)**, independent predictors of higher post-vaccination antibody levels were: previous SARS-CoV-2 infection (β=0.15 Scaled Luminescence units; 95%CI 0.06, 0.24), age <18 years (β=0.15; 95%CI, 0.05, 0.26), and receiving mRNA-vaccines (vs. a combination of mRNA and viral vector: β=0.23 for mRNA-1273 and β=0.10 for mRNA-BNT162b2). Lower antibody levels were associated with being over 6 months post-vaccination (vs. 0-3 months; β=-0.24; 95%CI -0.37, -0.12) and receiving the viral vector ChAdOx1-S vaccine for both doses (β=-0.46; 95%CI -0.64, -0.28). Biological sex, presence of immunosuppressive comorbidities, and interval between vaccine doses (3-8 weeks vs 9-16 weeks) were not independently associated with post-vaccination antibody levels. Similar associations were observed in stratified analyses investigating adults and children separately **(Figure S1).**

**Figure 6.**
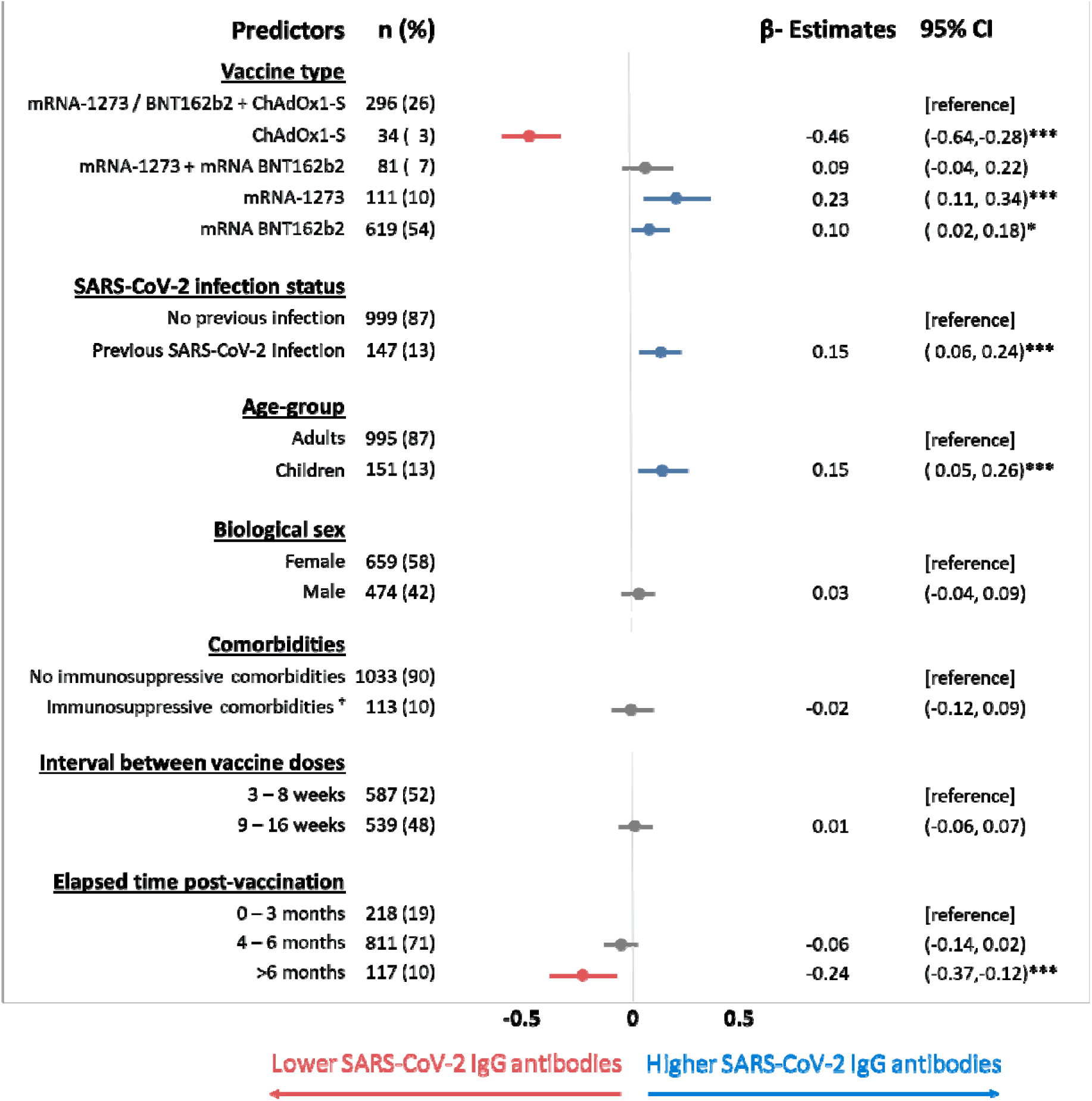
Predictors of anti-spike SARS-CoV-2 IgG antibody levels following two COVID-19 vaccine doses in the CHILD Cohort (n=1082). Cross-sectional serology data from dried blood spots collected between March 2021 and January 2022. Multivariable linear regression p-values ***≤0.001, **≤0.01, *≤0.05. ^†^Immunosuppressive comorbidities include current diagnosis of cancer, diabetes, pneumonia, and severe obesity.

## Interpretation

We report SARS-CoV-2 anti-spike IgG antibody responses among 3059 participants from a Canadian general population cohort of adults and children. Our cross-sectional analyses revealed an anti-spike IgG antibody seropositivity rate of 97% among 1146 double-vaccinated participants. Age, previous SARS-CoV-2 infection, vaccine type, and time since vaccination were identified as independent predictors of post-vaccination antibody levels.

The 97% seropositivity rate following two vaccine doses in our cohort is lower than rates reported in studies conducted in Italy (99.8%)^20^, Germany (99.8%)^22^ and England (98.9%)^23^. However, these studies included cohorts of healthcare workers who may have been exposed to repeated SARS-CoV-2 antigenic stimulation through prior infection, thereby influencing post-vaccination antibody levels with resultant higher seropositivity rates compared to our general population cohort.

Six percent of children and fourteen percent of adults in our cohort had hybrid immunity at the time of serologic testing (March 2021 to January 2022). Participants with hybrid immunity had significantly higher antibody levels compared to infection-naïve participants. While our study did not evaluate the chronological order of natural infection and vaccine-induced immunity, our data agrees with findings from cohorts of healthcare workers in Spain^30^, USA^36^ and Kuwait^37^ where individuals with prior COVID-19 had higher vaccine-induced antibody response.

SARS-CoV-2 anti-spike IgG antibody levels were lower in adults than children in our study, consistent with evidence in other studies that adults have reduced humoral responses to COVID-19 vaccine and other vaccine types compared to children^17–23^. We did not detect any sex differences, contrasting with other studies that found higher SARS-CoV-2 anti-spike IgG antibody levels in females compared to males^20,21,24,25^. However, similar to our findings, Brlić P. et al reported no significant gender-related differences in post-vaccination antibody levels among a cohort of 1072 healthcare workers in Croatia^26^. The lack of consistency regarding the association of biological sex and COVID-19 vaccine-induced humoral responses warrants further evaluation.

Participants in our cohort who received a combination of an mRNA and viral-vector vaccine had significantly higher antibody levels than homogenous viral-vector vaccinees, but lower levels than homogenous mRNA vaccine recipients. This contrasts with other findings that demonstrated higher antibody levels amongst heterogenous mRNA and viral-vector vaccinees compared to homogenous mRNA (BNT162b2) vaccine recipients^21,27–29^. Using a polynomial linear regression model to assess cross-sectional antibody levels, we showed that immune responses to mRNA-based vaccines were better maintained over the first six months compared to viral-vector vaccines. This is not surprising as mRNA vaccines induce more rapid humoral responses through short-lived plasma blasts compared to viral-vector vaccines^30^, in both plasma and saliva.^21^

SARS-CoV-2 anti-spike IgG antibody levels appear to wane three months after receipt of a second vaccine dose in our cohort. Similarly, a prospective general cohort study in Holland involving 2412 double-vaccinated participants showed a decline in antibody levels beginning at around four to six months post-vaccination^30^. Our findings on the timing of post-vaccination antibody decline provide further evidence supporting Health Canada’s current vaccination policy of a five-to-six-month interval between receipt of second vaccine dose and eligibility for third vaccine dose^31^.

We did not observe any differences between a 3 to 8 week dosing interval compared to a 9 to 16 week regimen, in contrast to data showing higher antibody levels after an extended regimen (6 to 14 weeks) compared with a shorter regimen (2 to 5 weeks)^53^. More research is required to understand and optimize COVID-19 vaccine dosing intervals.

### Limitations of the study

Our study has limitations. First, we did not capture the surge of new infections and re-infections by the more transmissible SARS-CoV-2 Omicron strain that emerged after the majority of our sample collection was complete. We expect re-infections to significantly impact the magnitude of post-vaccination antibody response. Second, the work presented here did not disaggregate the order of, and interval between infection and vaccination among study participants. Third, we relied on self-reported data (e.g., vaccine timing and type) which may introduce inaccuracies due to recall bias. Further, we did not measure other vaccine-induced immune responses, including T-cell, innate immune or memory-based responses, which may be important predictors of post-vaccination antibody response. Finally, our cohort over-represents higher socioeconomic status households which may limit generalizability to the Canadian general population.

## Conclusion

In this study involving over 3000 Canadians, including both children and adults and more than 1000 double-vaccinated individuals from the general population, we identified multiple health and demographic predictors of COVID-19 vaccine-induced antibody responses. Higher SARS-CoV-2 IgG antibody levels were associated with prior SARS-CoV-2 infection, recent vaccination, age under 18 years, and receiving mRNA vaccines, while no associations were observed for biological sex or time interval between doses. These findings help improve our understanding of COVID-19 induced humoral immune response in Canadian families and provide new information to inform future vaccine research and implementation strategies.

## Data Availability

All data produced in the present study are available upon reasonable request to the authors

https://childstudy.ca/for-researchers/study-data/

## Declarations

## Acknowledgements

We are grateful to all the families who took part in this CHILD COVID-19 Add-on Study and the entire CHILD Study team, which includes study site coordinators, research assistants, nurses, computer and laboratory technicians, clerical workers, research scientists, volunteers, managers, and receptionists at the following institutions: McMaster University, University of Manitoba, University of Alberta, SickKids, and University of British Columbia. We acknowledge Jay Onysko (Public Health Agency of Canada) who served on the CHILD COVID-19 Add-on Study Knowledge User Committee. We thank the main CHILD Cohort Study participant families for their dedication and commitment to advancing health research.

## Authors’ contributions

RA contributed to conceptualization, writing the original draft, formal analysis and reviewing and editing the final version. LL contributed to conceptualization, writing the original draft, project administration and supervision, formal analysis, and reviewing and editing the final version. GW and SG contributed to data curation and formal analysis. FB contributed to conceptualization, funding acquisition, data curation, formal analysis and reviewing and editing the final version. ML, CA, YG and MP contributed to serological analysis, editing and reviewing the final version. SB, JB, and DP, contributed to conceptualization, funding acquisition, and reviewing and editing the final version. ES, TJM, TES, PJM and PS contributed to conceptualization, funding acquisition, and reviewing and editing the final version. ND contributed to questionnaire review and revision, reviewing, and editing the final version. NR contributed to conceptualization, funding acquisition, project administration and reviewing the final version. MBA contributed to conceptualization, writing the original draft, funding acquisition, project administration and supervision, formal analysis, methodology and reviewing and editing the final version.

## Data availability

Existing CHILD researchers and CHILD COVID-19 Knowledge Users will be given rapid access to study data on request. New researchers interested in developing or collaborating on a project using CHILD data are encouraged to contact the study’s National Coordinating Centre for a formal request. Data request and access details are outlined on the CHILD website: https://childstudy.ca/for-researchers/study-data/.

## Ethics approval

This study was approved by Research ethics boards at the University of British Columbia (H20-02324), University of Alberta (Pro00102524), University of Manitoba (HS24250), The Hospital for Sick Children (1000071220) and McMaster University (1108).

## Supplementary Material

**Table S1.** Underlying medical conditions reported by double-vaccinated participants.

|  | Adults | Children |
| --- | --- | --- |
| Medical condition, n (%) | 995 (87) | 151 (13) |
| **Non-immunosuppressive conditions** |  |  |
| Allergies | 219 (22) | 34 (22) |
| Anxiety | 121 (12) | 17 (11) |
| Asthma/wheeze | 73 ( 7) | 13 ( 8) |
| Depression | 69 ( 7) | 4 ( 3) |
| Migraine headaches | 62 ( 6) | 3 ( 2) |
| Blood pressure, high | 61 ( 6) | 2 ( 1) |
| Chronic heartburn/reflux | 50 ( 5) | 1 ( 1) |
| Anemia (low blood count) | 40 ( 4) | 1 ( 1) |
| Irritable bowel syndrome | 32 ( 3) | 0 ( 0) |
| Cholesterol, high | 31 ( 3) | 0 ( 0) |
| Serious acne/skin problem | 24 ( 2) | 0 ( 0) |
| Attention deficit disorder (ADD,ADHD) | 17 ( 2) | 17 (11) |
| Urinary infections, recurrent | 9 ( 1) | 0 ( 0) |
| Learning disorder | 5 ( 1) | 6 ( 4) |
| Autism | 4 ( 0) | 6 ( 4) |
| Bronchitis | 4 ( 0) | 0 ( 0) |
| Heart disease | 4 ( 0) | 0 ( 0) |
| Osteoporosis | 4 ( 0) | 0 ( 0) |
| Behaviour problems | 3 ( 0) | 5 ( 3) |
| Bipolar disorder | 2 ( 0) | 0 ( 0) |
| Epilepsy | 2 ( 0) | 0 ( 0) |
| Croup | 1 ( 0) | 2 ( 1) |
| Stroke | 1 ( 0) | 0 ( 0) |
| Substance abuse (excluding alcohol) | 1 ( 0) | 0 ( 0) |
| **Immunosuppressive conditions<sup>†</sup>** |  |  |
| Arthritis | 40 ( 4) | 0 ( 0) |
| Immune disorder | 21 ( 2) | 2 ( 1) |
| Diabetes (non-insulin dependent) | 19 ( 2) | 0 ( 0) |
| Severe Obesity (BMI ≥40) | 16 ( 2) | 2 ( 1) |
| Inflammatory bowel diseases | 15 ( 2) | 0 ( 0) |
| Cancer | 8 ( 1) | 0 ( 0) |
| Kidney disease (including kidney stones) | 5 ( 1) | 1 ( 1) |
| Diabetes (insulin dependent) | 1 ( 0) | 1 ( 1) |
| Pneumonia | 1 ( 0) | 0 ( 0) |
| One or more immunosuppressive condition | 107 (11) | 6 ( 4) |
| **No medical condition** | 169 (17) | 50 (33) |

<sup>†</sup> Classified as immunosuppressive medical conditions or medical conditions requiring immunosuppressive therapy.

**Figure S1.**
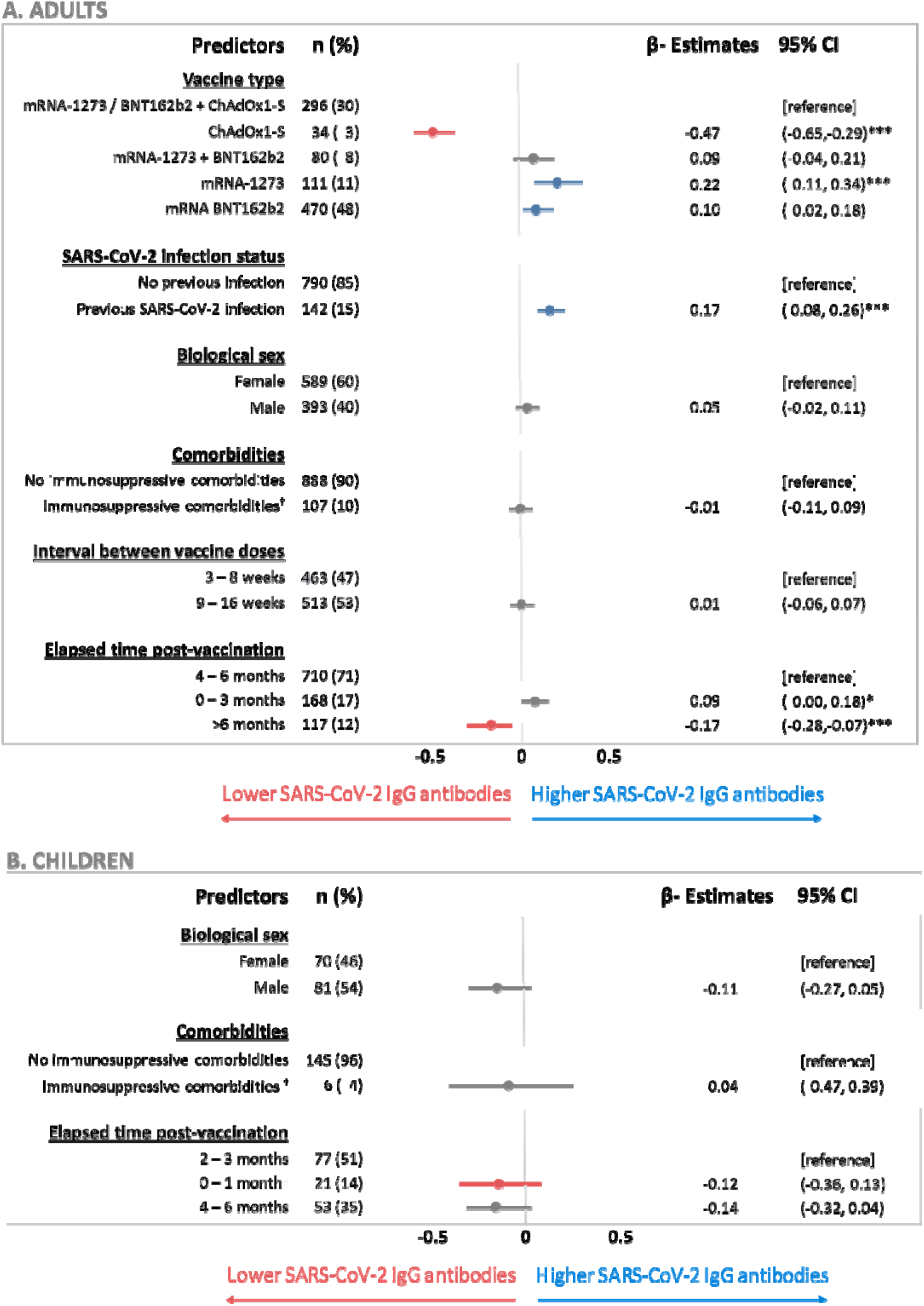
Multivariable linear regression: predictors of anti-spike IgG antibody levels following two COVID-19 vaccine doses in 932 adults (A) and 153 children (B). Cross-sectional serology data from dried blood spots collected between March 2021 and January 2022. *p*-value ***≤0.001, **≤0.01, *≤0.05. ^†^Immunosuppressive comorbidities include current diagnosis of cancer, diabetes, pneumonia, and severe obesity.

